# Fully Wearable Armband for Long-Term Peripheral Ultrasound Neuromodulation in Pain Management

**DOI:** 10.1101/2025.10.29.25339092

**Authors:** William D. Moscoso-Barrera, Yiming Han, Jinmo Jeong, QiLiang Chen, Thomas Wynn, Mengxia Yu, Kai Wing Kevin Tang, Mengmeng Yao, Ju-Chun Hsieh, David Wu, Landon Gauthreaux, Xiang Qian, Yaoyao Jia, Huiliang Wang

## Abstract

Chronic pain is a leading cause of medical consultations, often managed with opioids despite their high risk of addiction. Low-Intensity Focused Ultrasound (LIFU) neuromodulation has emerged as a promising non-invasive alternative for pain management. However, effective pain modulation often requires prolonged or continuous stimulation, and current LIFU devices are typically bulky and tethered to external power sources, limiting their applicability as wearable technologies for long-term use. This study introduces a fully wearable LIFU-based armband designed for peripheral nerve stimulation in pain management. It integrates a concave piezoelectric transducer, an acoustic hydrogel sheet for strong adhesion and extended use, as well as a miniaturized electronic circuit with a lithium-ion battery in a fabric armband for comfortable placement. LIFU was applied to the median nerve, and its effects on pain thresholds were evaluated in healthy volunteers using pressure algometry and cold pressor tasks. Results showed a significant increase in pain thresholds, with an average rise of 15%. These findings support the potential of wearable LIFU neuromodulation as a noninvasive and portable therapy for chronic pain management.

## Introduction

Chronic pain is a major public health issue that causes significant physical and psychological distress and is one of the most frequent reasons for hospital visits ^1^. In 2021, the Centers for Disease Control and Prevention (CDC) reported that approximately 21% of U.S. adults suffer from chronic pain ^2,3^, with neuropathic pain affecting 7–8% of the population (4). Neuropathic pain results from damage or disease affecting the somatosensory system ^4^ ^5^, including central neuronal injury in the brain or spinal cord, and peripheral nerve fiber damage, such as myelinated Aβ and Aδ fibers and unmyelinated C fibers. Current treatments—pharmacological, surgical, and neuromodulation-based—include nonsteroidal anti-inflammatory drugs (NSAIDs), opioids, and anticonvulsants ^6–8^, but these can lead to serious side effects such as overdose and addiction ^9^. Surgical and electrical stimulation therapies have shown effectiveness ^10,11^, although their invasiveness and long-term stability remain key challenges ^12–14^. Non-invasive, nerve-targeting therapies are still limited and often require administration in clinical settings. A wearable focused ultrasound (FUS) device may offer a promising solution for safe and effective pain management at home by non-invasively targeting peripheral nerves.

Low-intensity focused ultrasound (LIFU) has recently emerged as a new non-invasive neuromodulation treatment for pain. This technique delivers acoustic energy remotely to modulate the activity of both superficial and deep peripheral nerves in humans ^15–19^. Low-pressure ultrasound in a frequency range of 0.48–2.67 MHz can induce mechanical effects and provoke tactile and nociceptive responses that are reversible and harmless to surrounding tissues ^20^. Transient peripheral nerve stimulation with LIFU in healthy individuals could modulate cooling, warming, vibrotactile and pain sensations without increases in skin temperature, making it a potential non-invasive treatment modality for pain ^21,22^. However, the large size and complex design of the current LIFU devices greatly limit their adaptation as a wearable devices for long-term neuromodulation in pain management^23^.

In the present study, we introduce a novel fully wearable armband design called *PerNeUS* (**Per**ipheral **Ne**rve **U**ltrasound **S**timulator), which consists of a concave ultrasound transducer (CUT) and a bioadhesive acoustic hydrogel for ultrasound-based neuromodulation of peripheral nerves. The integrated system includes a miniaturized electronic circuit, a lithium-ion battery, and a fabric armband, enabling untethered operation and secure, comfortable placement during stimulation. To validate the wearable’s functionality, we conducted pressure algometry (PA) and cold pressor task (CPT) experiments in healthy volunteers, first using only the acoustic hydrogel integrated connected to a commercial ultrasound control system, and then using the acoustic hydrogel integrated connected to our own miniaturized circuits and battery as fully wearable device. Results demonstrated that LIFU stimulation significantly increased pain thresholds in healthy volunteers, suggesting an inhibitory effect on pain perception mediated by peripheral nerve ultrasound stimulation.

## Results

### Design and Characterization of the fully wearable armband

The design of the wearable armband consists of six components: a concave ultrasound transducer (CUT) (Lead zirconate titanate (PZT), DL-47, Del Piezo), an acoustic hydrogel, a 3D-printed polylactic acid (PLA) housing, a miniaturized electronic circuit, a lithium-ion battery, and a fabric armband. **(Fig. 1a-b)**. The miniaturized circuit and lithium-ion battery are housed in a dedicated pocket within the fabric armband. The CUT, acoustic hydrogel, and 3D-printed housing together form the transducer of the wearable, which is connected to the circuit embedded in the fabric armband (**Fig. 1c**). An acoustic hydrogel developed in our previous work ^24^ serves as a coupling medium between the concave structure of the wearable transducer and the skin, eliminating air gaps, ensuring efficient ultrasound transmission, and providing strong adhesion. The miniaturized electronic circuit is composed of three printed circuit boards (PCBs) powered by a battery, which together generate the LIFU signal delivered to the wearable transducer (**Fig. 1d**). Two daughterboards contain DC-DC converters that supply voltage to components on the motherboard. The motherboard includes a voltage regulator that powers both the oscillator and the microcontroller unit (MCU). These two components generate the modulated ultrasound signal, which is then amplified through a gate driver and a power amplifier. The fabrication process of the wearable transducer involves five steps, in which the three components are sequentially assembled **(Fig. 1e)**, with details provided in the Methods section. The wearable transducer is attached to the upper limb using a Velcro strap.

**Figure 1.**
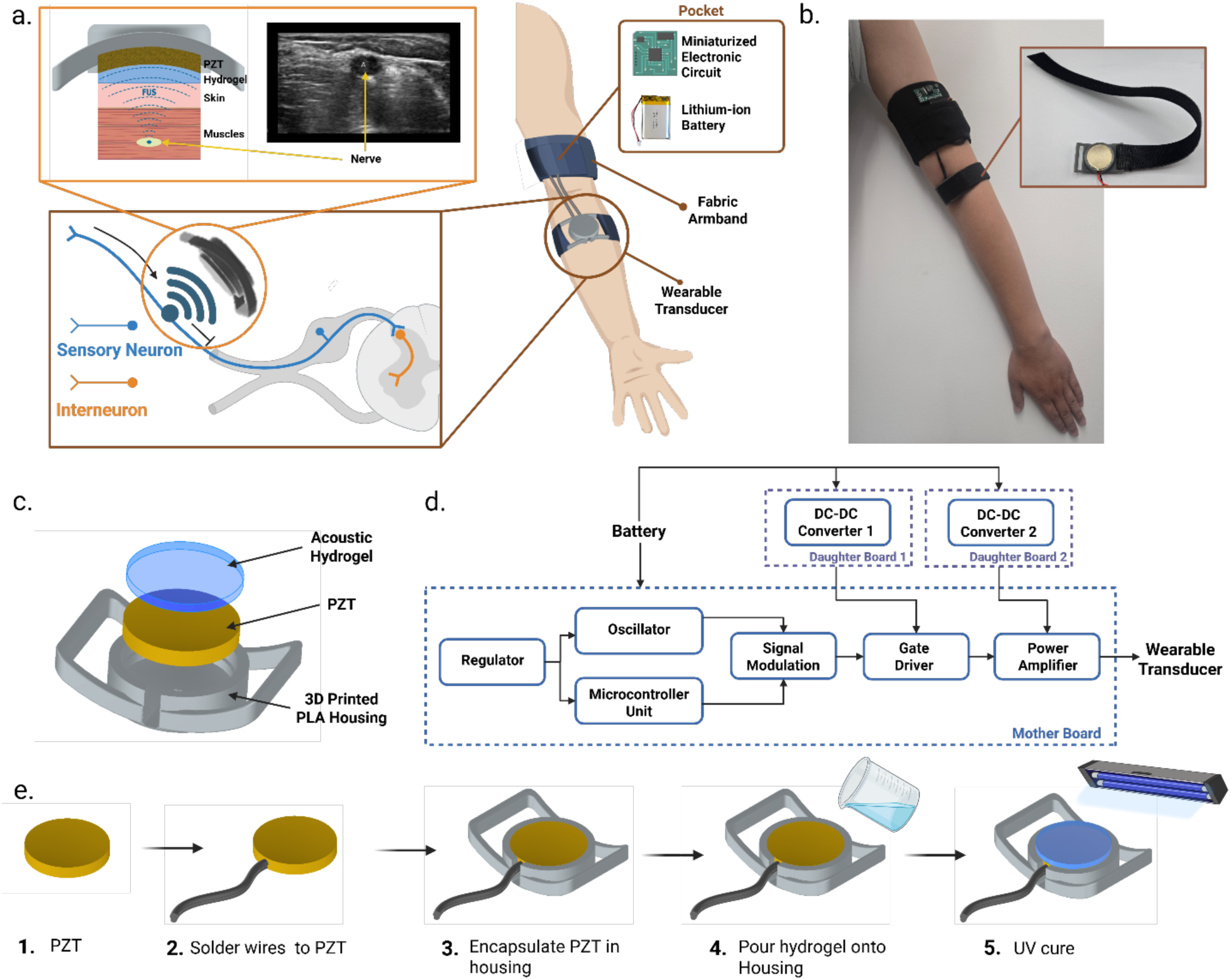
Fully wearable ultrasound device for peripheral neuromodulation. **(a)** Schematic of PerNeUS, a fully wearable FUS system for peripheral nerve stimulation to induce inhibitory modulation of sensory signaling for pain control. The upper panel shows a side view of the transducer’s layered stack and the FUS beam path to the target nerve, localized with diagnostic ultrasound. **(b)** Photograph of the complete system, which includes a fabric armband housing the miniaturized electronic circuitry and battery, and the wearable transducer; inset: Velcro-based attachment to the arm. **(c)** Schematic (exploded view) of the layered structure of the wearable transducer, comprising the concave PZT element and bioadhesive acoustic hydrogel, encapsulated in a compact 3D-printed housing. **(d)** Block diagram of the miniaturized electronics, consisting of a motherboard and two daughterboards. **(e)** Stepwise fabrication workflow of the wearable transducer.

An ultrasound frequency of 650 kHz PZT was utilized in the current study. The dimensions of the CUT have an outer diameter of 25 mm and a radius of curvature of 20 mm. The ultrasound properties of the CUT integrated with acoustic hydrogel were characterized in a deionized water tank **(Fig. 2a)**. With this geometric structure, the CUT can effectively focus the beam at distances ranging from 10 mm to 20 mm from the transducer **(Fig. 2b).** This focal depth range is sufficient for modulating peripheral nerves, including the median nerve ^25^. The measured full width at half maximum (FWHM) of the CUT integrated with acoustic hydrogel at this distance was 3.9 mm axially and 18.8 mm laterally **(Fig. 2c-d)**. The FWHM of the bare CUT was axially 3.5 mm and 18.5 mm laterally, showing a slight change but no significant difference of ultrasound beam profile between CUT with and without acoustic hydrogel **(Supplementary Fig. S1-3)**. These results confirm that hydrogel integration does not alter the focused beam characteristics. Electrical impedance tests were also conducted on the piezoelectric material, which showed that the resonance frequency corresponds to the lowest impedance of 28.84 ohms **(Supplementary Fig. S4).**

**Figure 2.**
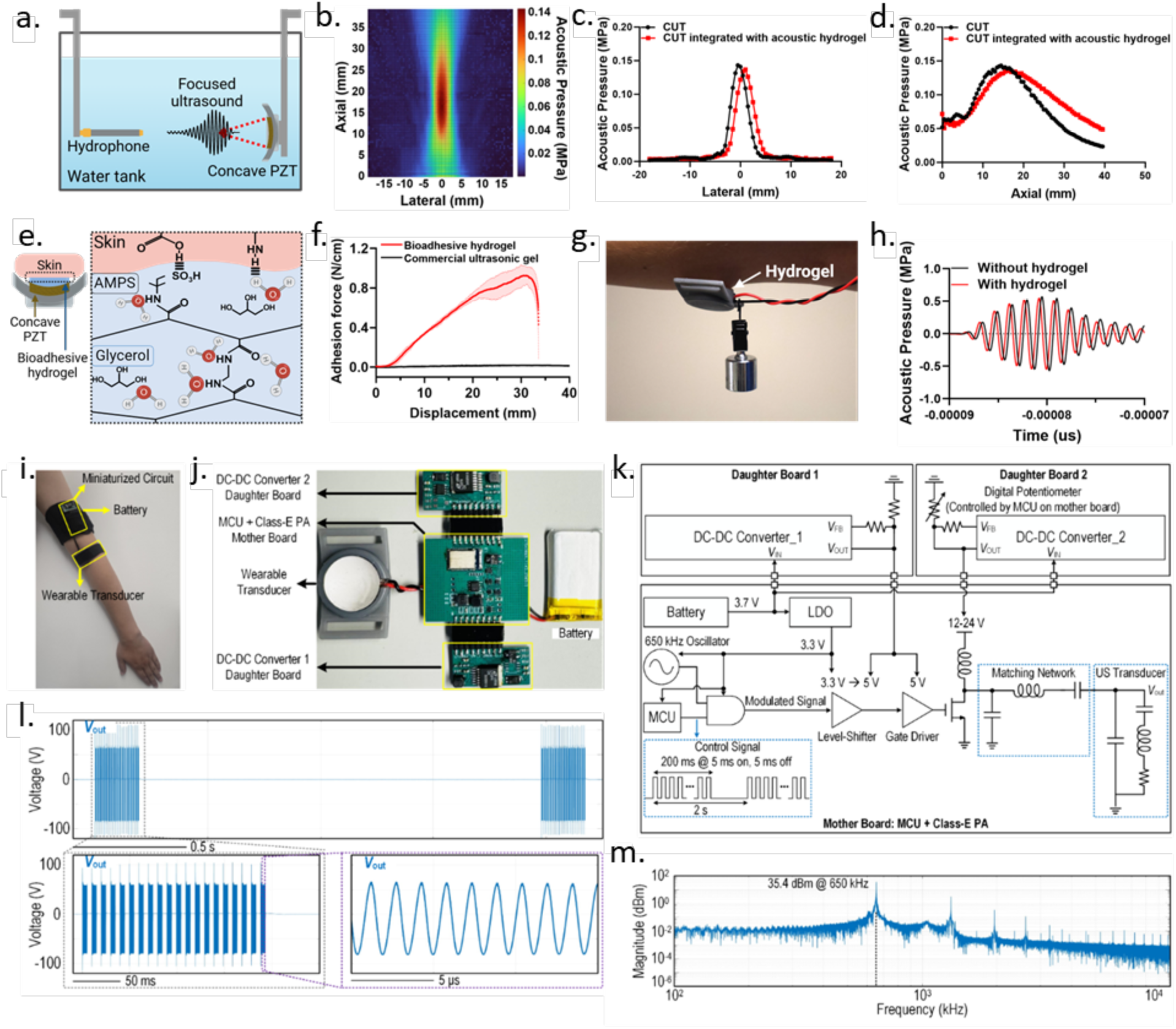
Characterization of the bioadhesive hydrogel integrated with the concave ultrasound transducer and the miniaturized electronic circuit. (**a)** Experimental setup for acoustic characterization of the concave ultrasound transducer (CUT). **(b)** Acoustic field distribution of the CUT integrated with acoustic hydrogel. **(c-d)** Comparison of the lateral and axial ultrasound beam profiles between CUT and CUT integrated with acoustic hydrogel in free-field. **(e)** Chemical structure of the bioadhesive hydrogel integrated with concave ultrasound transducer. **(f)** Adhesion force–displacement curves comparing the bioadhesive hydrogel and a commercial ultrasonic gel (Aquasonic 100, Parker) on skin (n=3). **(g)** Demonstration of adhesion strength: the hydrogel adhered to the underside of the forearm supports a 50 g weight. **(h)** Comparison of the acoustic pressure amplitude between ultrasound transducer with and without bioadhesive hydrogel layer. **(i)** Placement of the electronic circuit and battery within the wearable armband, and overview of the developed miniaturized circuit system. **(j)** Connection layout of components in the wearable circuit system. **(k)** Circuit schematic showing the implemented design with a motherboard and two daughterboards. **(l)** Measured transient output waveform of the electronic circuit connected to the transducer. **(m)** Frequency spectrum of the generated ultrasound output signal.

The acoustic hydrogel is composed of 2-acrylamido-2-methyl-1-propanesulfonic acid (AMPS) and glycerol, provides anti-dehydration properties and strong skin adhesion ^24^ **(Fig. 2e)**. Measured adhesion force-displacement curves of the acoustic hydrogel on skin were shown in **Fig. 2f**. The maximum adhesion force (0.931 ± 0.096 N/cm²) was sufficient for stable attachment during peripheral nerve stimulation, even before applying the fixation strap. In contrast, the maximum adhesion force of the commercial ultrasonic gel (Aquasonic 100, Parker) exhibited a maximum adhesion force of only 0.021 ± 0.002 N/cm², indicating negligible adhesion **(Fig. 2g; Supplementary Fig. S5)**. To evaluate the acoustic attenuation, the CUT device integrated with acoustic hydrogel was controlled by a commercial ultrasonic system (Vantage 64LE, Verasonics). The ultrasound intensity of the CUT reached approximately 0.57 MPa when 30 V was applied; with the hydrogel, the intensity was 0.54 MPa, representing only ∼5% attenuation (**Fig. 2h; Supplementary Fig. S6**).

We developed a compact, wireless, and wearable circuit system to drive ring-shaped ultrasound wearable transducer. The system (**Fig. 2i-j**) adopts a mother–daughter board architecture, powered by a 3.7 V, 550 mAh rechargeable lithium-polymer battery. The fully assembled board measures 4.3 × 4 cm², making it compact enough for seamless integration into a wearable armband. The motherboard houses the core driving circuitry, while two daughterboards include isolated DC-DC converters that supply different voltage levels to the motherboard. **Fig. 2k** illustrates the circuit implementation. The motherboard integrates a Class-E power amplifier (PoAm), microcontroller unit (MCU), low-dropout regulator (LDO), oscillator, and gate driver. The two DC-DC converters supply: (1) a fixed 5 V to the gate driver, and (2) an adjustable 12–24 V supply to the PoAm. The MCU wirelessly adjusts the PoAm voltage via a digital potentiometer. The LDO converts the 3.7 V battery input to 3.3 V to power the MCU and logic. A 3.3 V, 650 kHz square wave is generated by the oscillator, matching the center frequency of the ring-shaped transducer. The MCU modulates this waveform by gating the 650 kHz signal into bursts of 5 ms on and 5 ms off, repeated 20 times every 2 seconds. This modulated signal is level-shifted to 5 V and buffered by the gate driver, which in turn controls the gate of the Class-E PoAm. The PoAm, composed of an NMOS power transistor and a resonant matching network, efficiently amplifies and converts the modulated waveform into a duty-cycled 650 kHz sinewave, delivering 120–200 V peak-to-peak and up to 3.5 W of output power, with over 90% efficiency. This high-voltage, high-power output enables effective delivery of acoustic energy for peripheral nerve stimulation.

In addition, the ultrasound driver is wirelessly programmable via Bluetooth Low Energy (BLE). Users can adjust the PoAm supply voltage to modulate stimulation intensity and configure parameters such as timing, duration, and start/stop commands in real time. This wireless and compact design enables flexible, portable neuromodulation. **Fig. 2l** presents the measured transient output waveform of the driver circuit connected to the ring-shaped ultrasound transducer. The system delivers ultrasound stimulation every 2 seconds using a protocol consisting of 136 repetitions, each with a pulse repetition frequency (PRF) of 100 Hz and a burst duration (BD) of 200 ms. Within each burst, the waveform is modulated at a 50% duty cycle, corresponding to 5 ms on / 5 ms off intervals, repeated 20 times **(Supplementary Fig. S7)**. A zoomed-in view shows a clean sinusoidal waveform with a peak-to-peak amplitude of approximately 150 V. **Fig. 2m** displays the frequency spectrum of the output, with a strong spectral peak at 650 kHz and a magnitude of 35.4 dBm, corresponding to an estimated 3.5 W of AC output power—sufficient to drive the ring-shaped transducer effectively.

Using the ultrasound stimulation parameters with a 650 kHz wearable transducer, we performed nociceptive testing with pressure algometry (PoAm) and cold pressor task (CPT) in healthy human volunteers (n=8) to evaluate the changes in pain thresholds before and after LIFU stimulations (also referred to as Focused Ultrasound Stimulation - FUS) on the median nerve in the antecubital fossa **(Supplementary Fig. S8)**.

### Modulating pain thresholds in healthy volunteers with wearable

Neuromodulation has emerged as a promising field for pain treatment, with focused ultrasound (LIFU) showing significant potential in non-invasive neuromodulation. While most LIFU research for pain management focuses on brain stimulation, early studies demonstrated the feasibility of stimulating peripheral nerves using ultrasound. Gavrilov et al. showed that ultrasound pulses can evoke various sensations in humans, including tactile and pain responses ^15,16^. Subsequent studies explored the effects of intense focused ultrasound (iFU) on peripheral nerve sensitivity and characterized brain activity in response to peripheral nerve stimulation at different frequencies ^26^. Research has also investigated tactile and nociceptive responses to continuous and pulsed stimuli at varying frequencies ^19^. While limited in number and varying in ultrasound parameters, these studies provide a foundational framework for selecting ultrasound stimulation settings.

Here, we conducted seven experiments involving healthy volunteers. The first five experiments were performed by connecting only the wearable transducer connected to a commercial ultrasound control system **(Supplementary Fig. S9)**. This approach aimed to evaluate the effectiveness of focused ultrasound (FUS) stimulation on pain with our ultrasound transducer. The experiments included pressure algometry (PA), the cold pressor task (CPT), post-stimulation effects, and long-term acoustic hydrogel stability over a 21-day period. The remaining two experiments used the fully integrated wearable device to determine whether the custom-designed circuit could replicate the same neuromodulatory effects as the commercial system. These experiments also included pressure algometry and the Cold Pressor Task. All analyses from the seven experiments were conducted using normalized data.

### Pressure Algometry (PA)

The PA test measures pain detection thresholds (PDT) by applying controlled mechanical pressure to a specific point on the body. Using this test, we assess the effects of FUS neuromodulation on sensitivity to mechanically induced pain. Tests were conducted over three days, recording PDT data at three hand locations: wrist (P1), palm (P2), and dorsal middle finger (P3) **(Fig. 3a, Supplementary Fig. S10)**. On day one, baseline PDTs were recorded; on days two and three, LIFU and sham stimulations were administered in randomized order. The selected points (P1–P3) correspond to the anatomical pathway and branches of the median nerve in the hand ^27^. There were no significant differences in PDT between baseline and sham conditions **(Fig. 3b-i)**. Overall, P3 exhibited higher pain thresholds P1 and P2, as the nerve is in the middle finger and surrounded by minimal muscle and one of the phalangeal bones.

**Figure 3.**
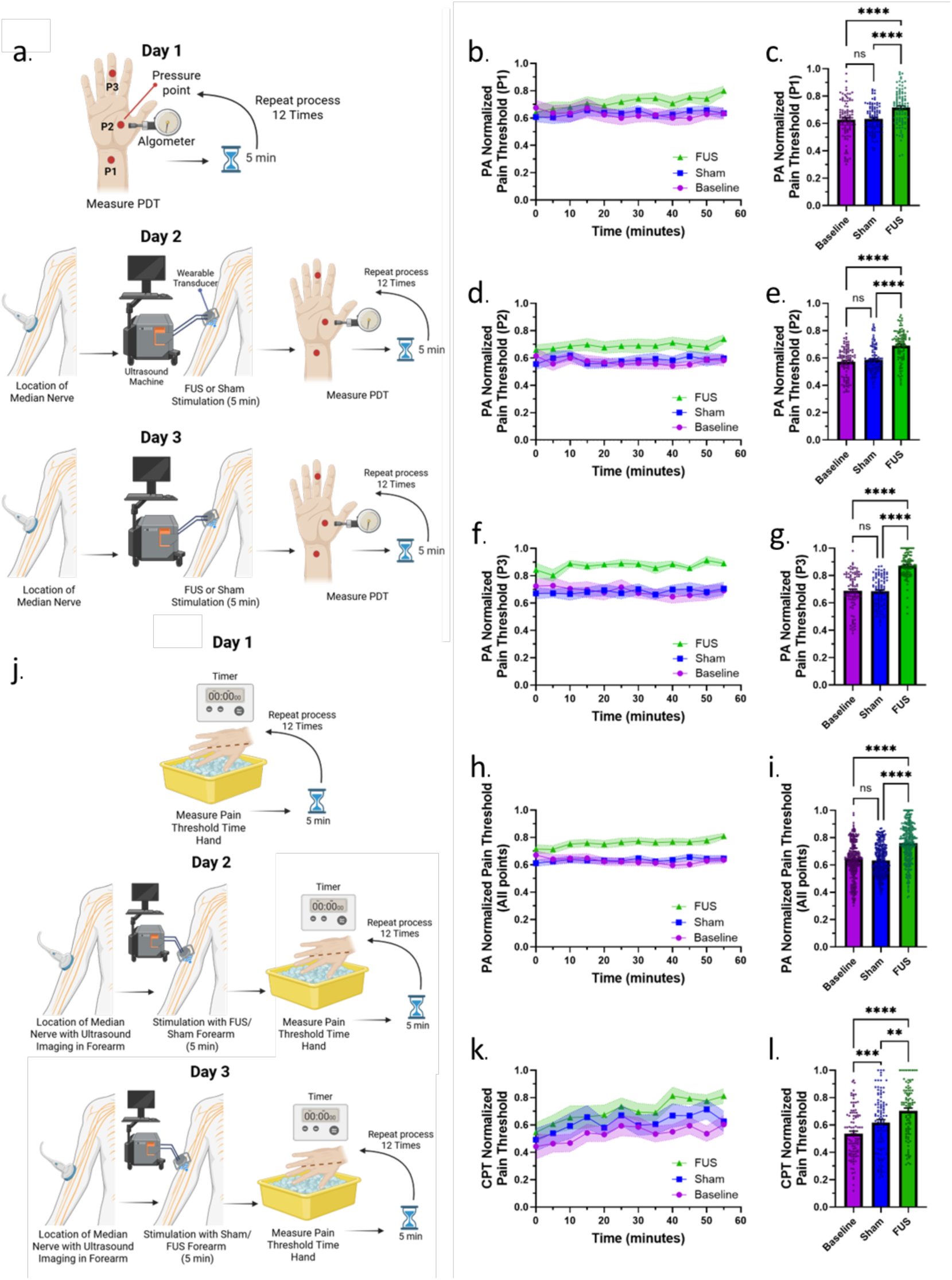
Pain Detection Thresholds tests using Pressure Algometry (PA) and Cold Pressor Task (CPT). **(a)** Experimental design diagram for the PA test. **(b, d, f)** Time-series plots showing aggregated normalized pain detection thresholds (PDTs) over time for sites P1, P2, and P3, respectively. **(c, e, g)** Aggregated normalized PDTs for individual measurement sites P1, P2, and P3. **(h)** Time-series plot of aggregated normalized PDTs across all measurement sites. **(i)** PA Aggregated normalized PDTs across all sites. **(j)** Experimental design diagram for the CPT test. **(k)** Time-series plot of aggregated normalized PDTs during the CPT test. **(l)** Aggregated normalized PDTs during the CPT test. Data are presented as mean ± s.e.m. unless otherwise indicated. Statistical analysis: PA and CPT tests (n = 8) analyzed using the Friedman test with Dunn’s multiple comparisons. Significance levels: *P < 0.05, **P < 0.01, *P < 0.001.

A non-parametric Friedman test was conducted to compare pressure pain thresholds across the three experimental conditions (baseline, sham, and FUS). The analysis, conducted in the experiment using the wearable transducer connected to the commercial ultrasound stimulation system, revealed a significant overall difference among the conditions across all measurement points (Friedman statistic = 213.1, Degrees of freedom or df = 2, p < 0.0001) **(Fig. 3i)**. Post hoc pairwise comparisons (Dunn’s test) indicated no significant difference between baseline and sham (p > 0.9999), whereas FUS stimulation significantly increased pain thresholds compared to both baseline and sham conditions (adjusted p < 0.0001). These findings demonstrate that FUS stimulation significantly increased pressure pain tolerance, as evidenced by higher PDTs across all tested sites. Time-series data confirm this trend: baseline and sham conditions remained similar, while PDTs following FUS were consistently elevated (**Fig. 3b, 3d, 3f, 3h**). Collectively, these results indicate that FUS exerts an inhibitory effect on pain transmission in the PA test.

### Cold Pressor Task (CPT)

The CPT was employed to evaluate tolerance to intense cold-induced nociception following FUS of the median nerve. The protocol was conducted over three consecutive days: baseline measurements were obtained on day 1, while FUS and sham stimulations were administered on days 2 and 3 in a randomized sequence **(Fig. 3j)**. A Friedman test revealed a significant overall difference in pain tolerance across conditions (Friedman statistic = 54.20, df = 2, p < 0.0001), indicating a measurable effect of FUS on cold pain response. This experiment was conducted using the wearable transducer connected to a commercial ultrasound stimulation system. Post hoc Dunn’s test (α = 0.05) showed a small but significant increase in tolerance between baseline and sham (adjusted p = 0.0003), reflecting a mild placebo effect. Importantly, FUS stimulation produced a further, statistically significant increase in pain tolerance compared to sham (p = 0.0031), with the strongest difference observed between baseline and FUS (p < 0.0001). These results confirm that FUS enhances cold pain tolerance in healthy individuals, likely through inhibitory modulation of peripheral sensory nerve activity that decreases nociceptive signaling (**Fig. 3k–l**).

### Post-stimulation effects of FUS on peripheral pain control

To evaluate the duration of the observed analgesic effects, two additional experiments—one with PA and one with CPT—were conducted. The objective was to determine the persistence of pain inhibition following FUS stimulation. By analyzing the duration of stimulation, we aim to determine the optimal number of stimulations that could be applied in future studies for pain treatment. The PA tests involved four healthy volunteers and were conducted over two days using the wearable transducer connected to a commercial ultrasound system **(Fig. 4a)**. Baseline PDTs were recorded on day one, and stimulation was administered on day two. Pain thresholds were measured hourly for nine hours following stimulation. Results indicated that the FUS-induced increase in pain threshold lasted approximately 8–9 hours, after which PDTs returned to baseline (**Fig. 4b**). Comparison between baseline and post-stimulation PPTs showed a statistically significant difference, confirmed by a Wilcoxon matched-pairs signed-rank test (p = 0.0020) **(Fig. 4c)**. The same experiment was conducted using the Cold Pressor Task (CPT) over a 9-hour period with four healthy volunteers. Data was collected every hour across two consecutive days: baseline measurements were taken on day one, and post-stimulation measurements were taken on day two following FUS stimulation **(Fig. 4d)**. A significant difference between baseline and post-stimulation conditions was observed (Wilcoxon p = 0.0020) **(Fig. 4f)**. Unlike the PA test, the CPT results suggested that the analgesic effect persisted beyond 9 hours, as participants maintained longer hand immersion in ice water (**Fig. 4e**). These results indicate that FUS may produce sustained pain inhibition lasting more than 9 hours.

**Figure 4.**
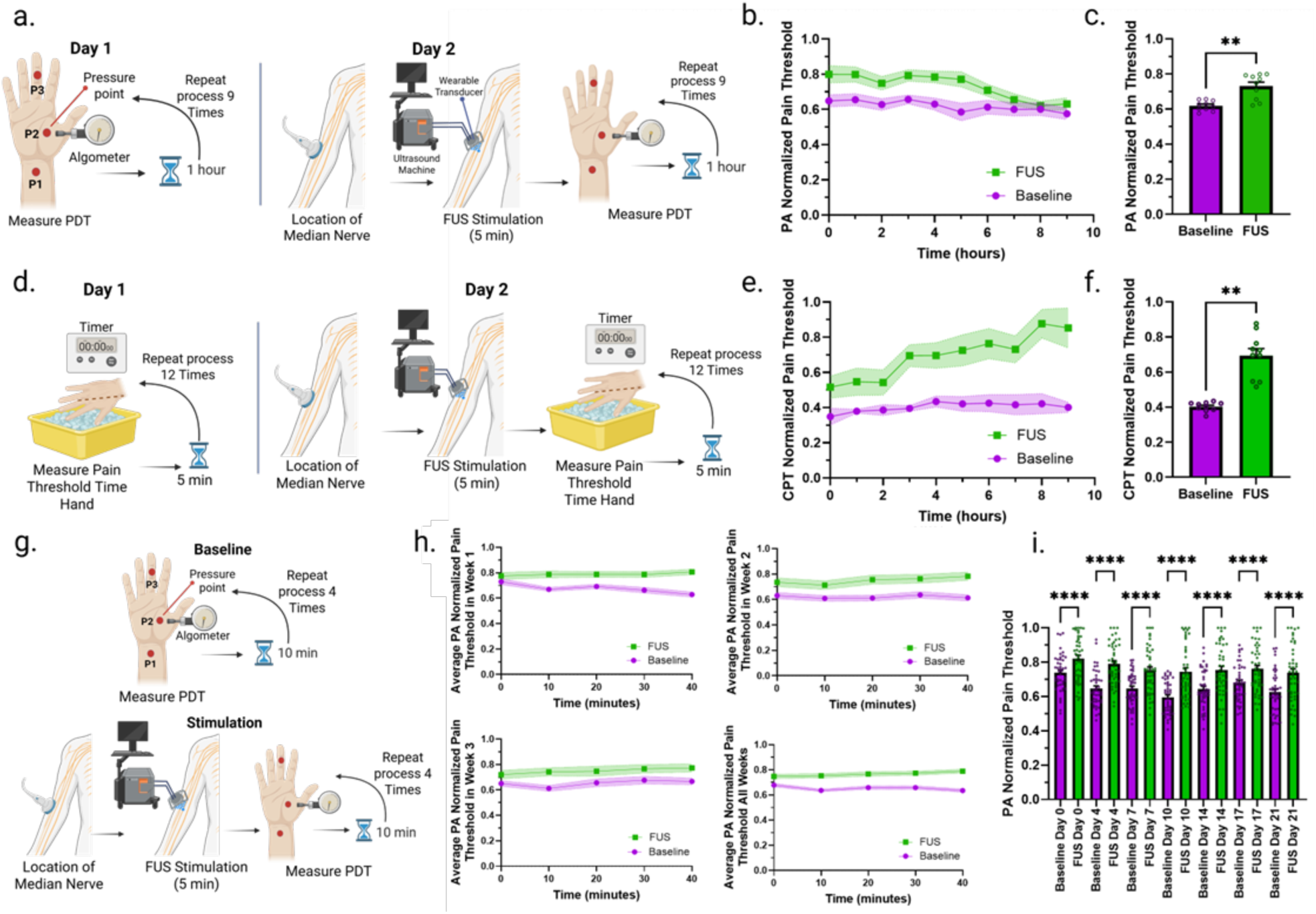
Post-stimulation effects of focused ultrasound (FUS) and long-term stability tests in pain management. **(a)** Experimental design diagram for the 9-hour pain-measurement test using Pressure Algometry (PA). **(b)** Time-series plots of aggregated normalized pain detection thresholds (PDTs) by experimental condition across all measurement sites, showing FUS-induced changes over the 9-hour period. **(c)** PA Aggregated normalized PDTs by experimental condition across all measurement sites, illustrating FUS-related effects over 9 hours. **(d)** Experimental design diagram for the 9-hour pain-measurement test using the Cold Pressor Task (CPT). **(e)** Time-series plots of aggregated normalized PDTs during the CPT test, showing FUS-induced changes over 9 hours. **(f)** Aggregated normalized PDTs during the CPT test, demonstrating sustained FUS effects over 9 hours. **(g)** Experimental design diagram for the 21-day acoustic- hydrogel durability test. **(h)** Time-series plots of aggregated normalized PDTs by experimental condition across all measurement sites over 21 days of repeated use. **(i)** Aggregated normalized PDTs measured every 3 days by experimental condition across all sites, illustrating long-term stability of the bioadhesive hydrogel and FUS efficacy. Data are presented as mean ± s.e.m. unless otherwise indicated. Statistical analysis: PA (9 h, n = 4), CPT (9 h, n = 3), and 21-day (n = 3) tests were analyzed using the Friedman test with Dunn’s multiple comparisons. Significance levels: *P < 0.05, **P < 0.01, *P < 0.001.

### Long-term stability of wearable transducer in pain management

The durability and functionality of the acoustic hydrogel were evaluated after repeated use for median nerve stimulation. This test assessed the feasibility of reusing the same hydrogel for extended periods, confirming the practicality of the wearable for long-term pain management. Experiments were conducted over 21 days, with measurements taken every three days. On each session, baseline PDTs were recorded, followed by LIFU stimulation and post-stimulation PDT measurements (**Fig. 4g**). It was confirmed that the wearable designed in this study, used for three weeks with the same acoustic hydrogel, successfully stimulated and generated an increase in pain threshold, showing significant differences between baseline and FUS in each measurement over the 21-day period (Dunn’s multiple comparisons test, α = 0.05, day 0 p = 0.0081, days 3, 7, 10, 14 and 21 p < 0.0001, day 17 p = 0.0009) **(Fig. 4h-i)**. Hydrogel stability testing followed the previously published protocol ^24^. **Supplementary Fig. S11** shows the hydrogel’s physical condition over 21 days, exhibiting no visible degradation and maintaining stable acoustic coupling properties for transmitting FUS energy to the tissue.

### *PerNeUS* Device Testing for Pain Management

Finally, the fully integrated wearable armband (*PerNeUS*) with miniaturized circuits was evaluated using both PA and CPT tests to assess its efficacy in modulating pain detection thresholds. Each test involved four healthy volunteers. The PA test was performed over three days. On day one, baseline measurements were collected; on days two and three, participants received either LIFU or sham stimulation (**Fig. 5a**). PDTs were measured at the same anatomical points as in previous experiments. Detailed data for each point (P1–P3), including normalized PDTs and bar plots, are shown in **Supplementary Fig. S12**. Dunn’s multiple comparisons test (α = 0.05) revealed no significant difference between baseline and sham (p > 0.9999) (**Fig. 5c**). However, LIFU stimulation significantly increased PDTs compared to both baseline and sham (p < 0.0001), with consistently elevated thresholds throughout the experiment (**Fig. 5d**). Similarly, the CPT test spanned three days—baseline, sham, and FUS conditions (**Fig. 5b**). While initial PDT levels were comparable, FUS stimulation produced a clear increase in cold pain tolerance relative to baseline and sham (**Fig. 5e**). No significant difference was observed between baseline and sham (p > 0.9999), but a significant difference emerged between FUS and baseline (p < 0.0001) (**Fig. 5f**).

**Figure 5.**
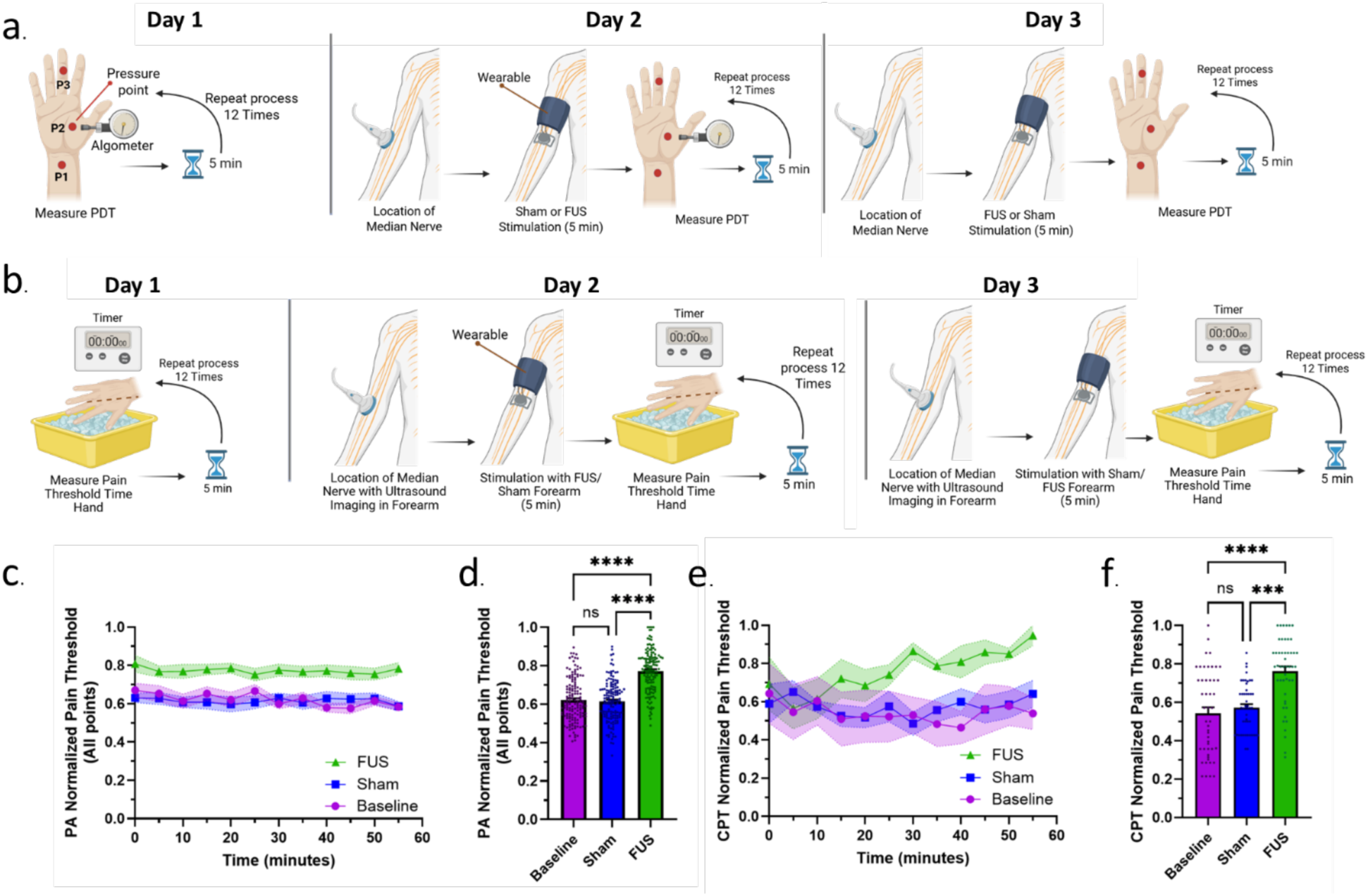
Pain Detection Threshold tests using the fully wearable armband device with Pressure Algometry (PA) and Cold Pressor Task (CPT). **(a)** Experimental design diagram for the PA test using the fully wearable device. **(b)** Experimental design diagram for the CPT test using the fully wearable device. **(c)** Time-series plots of aggregated normalized pain detection thresholds (PDTs) across all measurement sites during the PA test. **(d)** Aggregated normalized PDTs across all measurement sites during the PA test. **(e)** Time-series plots of aggregated normalized PDTs during the CPT test. **(f)** Aggregated normalized PDTs during the CPT test. The fully wearable device integrates both the ultrasound transducer and the miniaturized electronic circuit within a fabric armband. Data are presented as mean ± s.e.m. unless otherwise indicated. Statistical analysis: PA test (n = 4) and CPT test (n = 4) were analyzed using the Friedman test with Dunn’s multiple comparisons. Significance levels: *P < 0.05, **P < 0.01, *P < 0.001.

These findings confirm that the fully wearable system effectively induces pain inhibition, comparable to results obtained with the commercial ultrasound system (**Supplementary Fig. S13**). This demonstrates that *PerNeUS* delivers reliable and sustained ultrasound neuromodulation for pain management in a fully wearable format.

## Discussion

In 1965, Melzack and Wall proposed the Gate Control Theory of Pain, describing a mechanism through which painful sensations can be attenuated or inhibited by activating non-painful sensory inputs ^28^. Pain signals are transmitted from the periphery to the central nervous system (CNS) via myelinated A-delta fibers or unmyelinated C fibers in response to pressure, temperature, or injury stimuli.

Ultrasound stimulation of peripheral nerves operates through mechano-transduction, where ultrasound waves generate mechanical stimuli that induces the opening of mechanosensitive ion channels in nerve fibers. These include TRP channels (TRPV1, TRPA1), ASICs (acid-sensing ion channels), and Piezo channels, which are essential for converting mechanical stimuli into electrical signals i.e., neural signal ^29^. We believe that ultrasound stimulation can induce localized mechanical deformation, activate these ion channels and altering neuronal firing patterns and hence modulating the peripheral nerve activity. Additionally, previous studies have shown that ultrasound has a preferential effect on larger Aβ fibers, likely due to their higher mechanical threshold compared to smaller C fibers. This selective activation is particularly relevant for pain management, as Aβ fiber stimulation, according to the Gate Control Theory, inhibits pain transmission at the spinal cord level, thereby reducing pain perception ^30^. Moreover, ultrasound-induced changes in nerve excitability have been associated with a reduction in ectopic discharges in C fibers, which are strongly linked to chronic and neuropathic pain ^31^. This suggests that ultrasound neuromodulation may predominantly exert inhibitory effects ^32,33^. By modulating the excitability of peripheral nerve fibers and inhibiting the propagation of aberrant discharges, ultrasound effectively reduces pathological pain signaling that contributes to pain persistence. Taken together, these phenomena highlight the potential of ultrasound to reverse or prevent maladaptive neuroplastic changes associated with chronic pain, offering an innovative therapeutic approach for conditions such as neuropathic pain, where conventional pain management strategies often fall short.

We present a novel fully wearable device, named *PerNeUS* that integrates a concave ultrasound transducer (CUT), a bioadhesive acoustic interface for long-term use, a miniaturized electronic circuit, and battery embedded in a fabric armband specifically designed for peripheral pain management. Our study demonstrates that this fully wearable system enables noninvasive stimulation of peripheral nerves—here, the median nerve. The median nerve, like other peripheral nerves targeted for pain control, is located approximately 1.5 to 2 cm beneath the skin, making it accessible to a single CUT. This geometry is advantageous for wearable applications, as it permits the use of a single lightweight element integrated into a fully wearable system (total weight 93 g). However, CUTs have limitations: their concave structure creates a significant air gap when placed on the skin, leading to acoustic impedance mismatch and reduced ultrasound transmission ^34^. While commercial ultrasound gels can mitigate this, they dehydrate rapidly and are impractical for long-term neuromodulation ^35^. Therefore, integrating a bioadhesive hydrogel into the wearable design is essential to fill the air gap, maintain skin contact, and ensure consistent subcutaneous ultrasound delivery.

We selected parameters based on prior studies reporting effective peripheral nerve modulation using frequencies between 300 and 900 kHz ^19^. Pulse durations ranging from 5 ms to 500 ms have successfully elicited sensations in humans, as shown by Gavrilov ^15^. In our case, we used a pulse duration of 200 ms and a burst repetition frequency of 100 Hz ^19^, as demonstrated by Riis and Kubanek to be effective for human peripheral nerve stimulation. The spatial-peak pulse-average intensity (Isppa) was measured at 41.4 W/cm², well below the FDA’s recommended safety limit of 190 W/cm², ensuring biosafety compliance^19^.

Through Pressure Algometry and Cold Pressor Task tests, we validated that neurostimulation with specific LIFU parameters can increase the pain threshold, indicating inhibitory effects on nociceptive signaling. These findings align with the Gate Control Theory of Pain, as non-painful stimuli delivered via LIFU produced measurable effects on nerve fibers in the tests performed. In the Pressure Algometry test, which primarily probes A-delta fibers responsible for transmitting rapid, acute pain, LIFU neuromodulation effectively enhanced the capacity to withstand controlled mechanical pressure at three specific points along the median nerve pathway. Similarly, in the CPT, which engages A-delta and C fibers due to the prolonged nature of the intense cold stimulus, participants demonstrated an increased tolerance to the stimulus, reflected in a longer exposure time following LIFU stimulation. We also compared the wearable transducer driven by a commercial system (gold standard) with the fully wearable device during PA and CPT. The results showed that pain thresholds increased similarly in both cases, demonstrating the effectiveness of our miniaturized, wearable circuit design. These two tests collectively highlight the potential application of *PerNeUS* device as a promising tool for analgesic treatments. Additionally, we confirmed that this effect can last between 8 and 9 hours, suggesting feasibility of 2–3 daily sessions with only a few minutes of stimulation. The study also demonstrated the feasibility of long-term use of the wearable device facilitated by the acoustic hydrogel, which could simplify the administration of pain therapies. In a previous study, we developed and validated the acoustic hydrogel used in this device. It exhibited an acoustic impedance of 2.17 MRayl ^24^, closely matching that of human skin (1.99 MRayl) ^36^, which enables efficient ultrasound transmission. Its composition of AMPS and glycerol provides dehydration resistance and strong skin adhesion, making it ideal for prolonged coupling in wearable systems^24^.

The inhibitory effects of pain observed in our tests are consistent with previous studies on neurostimulation. Although limited research has been conducted on humans using LIFU, similar effects on peripheral nerves have been demonstrated with electrical stimulation. Animal studies have provided evidence of pain reduction through neurostimulation. For instance, in a study using a cat model, stimulation of the sciatic and posterior tibial nerves reduced C-fiber pain response at the spinal cord level ^37^. Additionally, analgesic effects have been achieved in rats through stimulation of nerves such as the sciatic nerve ^38,39^. Several studies have been conducted in humans using electrical stimulation of specific nerves, including the median, ulnar, sciatic, genitofemoral, and vagus nerves, demonstrating potential for chronic pain treatment ^40–43^. One such study focused on median nerve stimulation, which led to a reduction in neuropathic pain following traumatic amputation and reimplantation of the index and middle fingers ^44^. Some animal studies have been reported regarding the use of focused ultrasound (FUS). One study examined the effects of sciatic nerve stimulation in a frog model, reporting FUS-induced inhibition attributed to conduction block ^17^. Another study demonstrated FUS effects on the left cervical vagus nerve conduction in Long Evans rats, observing vagus nerve inhibition in most cases ^45^. Similar results were found in a study applying FUS to the sciatic nerve in mice, resulting in temporary blocks of peripheral nerves involved in pain signaling ^46^.

Regarding long-term pain inhibition, we believe that FUS preferentially recruits non-nociceptive A-beta fibers, sustainedly activating inhibitory interneurons and thereby reducing the conduction and firing of nociceptive A-delta and C fibers when pain stimuli occur. Studies on cortical neurons have shown that FUS mechanically interacts with the cell, activating specific mechanosensitive ion channels that are permeable to calcium. This activation leads to a gradual accumulation of calcium ions, which can trigger the opening of calcium-sensitive sodium channels. As a result, the cell membrane depolarizes, further activating voltage-gated calcium channels and modulating neuronal excitability ^47^. Based on this mechanism, transcranial ultrasound stimulation (TUS) studies have observed a significant increase in the functional connectivity of the dorsal anterior cingulate cortex (dACC) in deep cortical regions of humans for at least 50 minutes after stimulation ^48^. This convergence of calcium signaling and lasting functional effects suggests that FUS may induce long-term neuroplasticity. A systematic review by Zheng et al. concluded that repetitive neuromodulation is generally effective as a long-term complementary treatment for chronic pain, reinforcing the need to develop portable technologies suited for extended use ^49^.

Another key aspect of *PerNeUS* is its compact size and wearability, which make it highly practical for real-world applications. For example, it could be used for pain management in non-hospital settings, serving as a useful tool for physicians in outpatient clinics as well as for at-home treatments. Moreover, its operation via a lithium-ion battery makes it suitable for environments where rapid pain relief is needed but electrical infrastructure is not readily available. While several studies have focused on the miniaturization of ultrasound circuits ^50^, most have been oriented towards imaging applications or the external activation of electrical stimulation systems ^51^. The challenge of low-intensity focused ultrasound (LIFU) stimulation lies in generating pulsed waveforms with peak-to-peak voltages exceeding 100 V, which are necessary to deliver sufficient acoustic power (>1 Watts) to elicit neuromodulatory effects. To date, some research efforts have managed to generate waveforms reaching ∼60 V peak-to-peak, but with output power levels below 1 W ^52,53^. Consequently, commercially available systems for focused ultrasound stimulation remain bulky, relying on large-scale electronics, and are thus limited to clinical or hospital settings, lacking portability. Through a miniaturized architecture—with a main board housing the microcontroller, oscillator, and power amplifier, and two daughter boards providing high-voltage rails to specific stages—we generated output signals of 120–200 V peak-to-peak and ∼3.5 W of power. This compact design enabled, for the first time, the development of a fully portable and wearable focused ultrasound device, broadening the potential for noninvasive neuromodulation in pain management.

We anticipate that *PerNeUS* will enable future treatment of neuropathic conditions such as carpal tunnel syndrome, cubital tunnel syndrome, radial nerve palsy, rheumatoid arthritis, and diabetic neuropathy, among others. These neuropathies often result in chronic pain, with current treatments providing initial relief but frequently leading to symptom recurrence. Another significant concern in treating these conditions is the reliance on opioid therapy when initial treatments fail to alleviate pain, given the severe side effects associated with opioids, including addiction and overdose ^54–57^. Thus, our technology offers a promising pathway for innovative, noninvasive pain treatments that reduce dependence on pharmacological interventions. Implementing this technology could facilitate pain treatment beyond clinical settings, enabling at-home therapy and potentially reducing the need for invasive procedures, surgery, or opioids. Increased adoption of non-invasive technologies could empower more clinicians and patients to access peripheral nerve stimulation. In the future, patients may even self-administer therapy using portable devices, similar to existing electrical stimulation systems or TENS units ^58^.

This study has certain limitations, primarily due to the small sample size of healthy volunteers. Future steps include conducting these tests on larger cohorts of participants. Following these tests, preliminary clinical trials could be conducted with patient groups suffering from the conditions, evaluating the effects of LIFU for pain treatment with 2-3 daily stimulations over several days. It is also recommended that these studies incorporate pain scales, questionnaires, and sleep quality surveys. Lastly, it will be essential to assess the durability of the pain treatment after completion, evaluating pain perception in the weeks following treatment to establish the optimal frequency of application.

## Methods

### Materials and fabrication of hydrogel

The preparation of the bioadhesive hydrogel began by preparing the hydrogel solution. First, AMPS (Sigma-Aldrich) was dissolved in deionized (DI) water at a 1:1 ratio using a vortex mixer for 30s. Subsequently, glycerol (Alfa Aesar) with 20 wt% relative to the total solution was added to the AMPS/DI water mixture using a vortex mixer for 30 seconds. N, N’-Methylenebis(acrylamide) (MBAA crosslinker, Sigma-Aldrich) at ∼0.16 wt% was then added and mixed for 60 seconds. Irgacure 2959 (2-Hydroxy-4’-(2- hydroxyethoxy)-2-methylpropiophenone 98%, Sigma-Aldrich) at ∼0.59 wt%, as the photoinitiator, was mixed for 30 s. The solution was stirred additionally for 30 minutes ^24^.

#### Adhesion force of the bioadhesive hydrogel with skin

The adhesion force was measured using a 90° peel tester (FB5, Torbal) according to ASTM F2255-05 and ASTM F2256-05 standards ^24^. The sample size was 20 × 50 × 2 mm³ (width × length × thickness), with a Kapton film attached to the back to prevent stretching during peeling. Adhesion between the skin and hydrogel was measured by by applying the sample to skin and peeling at 90° at 68 mm/min.

### Fabrication of the Fully Wearable *PerNeUS* Device

The fabrication of the fully wearable *PerNeUs* device was carried out in three stages. In the first stage, the wearable transducer was constructed as shown in **Fig. 1e**. A piezoelectric material (DL-47, Del Piezo) was used, and wires were soldered to establish the electrical connection to the circuit. The transducer was then encapsulated within a housing using epoxy (Epoxy Instant Mix, Loctite). The prepared acoustic hydrogel was subsequently placed onto the concave aperture and cured under UV light for 15 minutes. A Velcro strap was sewn to the wearable to allow secure attachment to the arm. The second stage involved the fabrication of the electronic boards, where components were soldered onto both the main and two daughter boards (**Fig. 2b**), followed by the attachment of connectors for interfacing with the transducer and battery. In the third stage, a fabric armband was made from elastic textile material, incorporating a pocket for the placement of the electronic boards and battery. Finally, all components (boards, battery, and transducer) were assembled inside the fabric armband and interconnected using wires (**Fig. 1b**).

### Acoustic characterization

Acoustic properties of the concave ultrasound transducer were measured in a deionized (DI) water tank using a needle hydrophone (HNR-0500, Onda) mounted on a three-axis stage and connected to an oscilloscope (SDS 1204-XE, Siglent). Ultrasound was generated by a commercial ultrasonic system (Vantage 65LE, Verasonics), and the ultrasound paradigm was 650 kHz center frequency, 100 Hz pulse repetition frequency (PRF), 200 ms pulse duration (PD, 50% duty cycle (5 ms ON)). Acoustic field distribution was mapped in 500 µm increments (0 – 40 mm from the transducer) over a 38 × 38 mm grid, with control and signal processing performed using a custom MATLAB **®** program. The spatial-peak pulse average (Isppa) was calculated by the following equations ^59^:

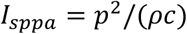

Where p is the acoustic pressure, 𝜌 is the density of the deionized water (1000 kg/m^3^), and 𝑐 is the speed of sound of the deionized water (1480 m/s).

When measuring the acoustic properties of the concave ultrasound transducer integrated with bioadhesive hydrogel, the hydrogel was wrapped with a thin latex layer to prevent swelling due to contact with water in the deionized tank.

### Ultrasound Parameters for Peripheral Neuromodulation

For the generation of low-intensity focused ultrasound (LIFU) in the tests using a commercial system connected to the wearable transducer, we employed the image-guided therapy ultrasound system (BBBoq, Image Guided Therapy System) **(Supplementary Fig. S9)**. A custom code was developed to generate the ultrasound protocol using the Arduino® Integrated Development Environment (IDE) and implemented on an Arduino Mega 2560 board. The board generates pulses that are sent to the trigger input of the commercial ultrasound system, configured for activation on a rising edge. Upon receiving the signal from the Arduino, the system emits a 5-millisecond ultrasound pulse directed to the wearable transducer. The procedure used a transducer operating at 650 kHz fundamental frequency, delivering ultrasound waves with a burst duration (BD) of 200 ms, burst repetition frequency (BRF) of 100 Hz, and an inter-stimulus interval (ISI) of 2 s. This protocol was repeated for a total sonification duration (SD) of 5 min. In the fully wearable device, custom firmware on the microcontroller, together with a 650 kHz square-wave oscillator, replicated the same waveform parameters used with the commercial system.

### Pain Threshold Tests

#### Pressure Algometry (PA) Tests

In the first test (commercial ultrasound system connected to wearable transducer), we measured the pain detection threshold (PDT) using pressure algometry of the dominant hand of eight healthy volunteers (5 males and 3 females, aged 21-39 with a mean age of 30.8 ± 6.3 years).

PA was performed with the Wagner FPX-25 (Wagner Instruments, Greenwich, GT). This test was conducted in three sessions on different days **(Fig. 3a)**. On day 1, baseline PDTs were recorded at three median-nerve dermatome sites: the palmar wrist, the middle of the palm, and the dorsal middle finger. PDT measurements were performed over 1 h with 5-min intervals (**Supplementary Fig. S10**). On days 2 and 3, the nerve was first located using a portable ultrasound (**Supplementary Fig. S8**). Participants served as their own controls, and counterbalancing minimized order effects: one subgroup received LIFU on day 2 and sham on day 3, and the other subgroup received sham on day 2 and LIFU on day 3.

In the sixth test, the *PerNeUS* device was used to measure PDTs using the PA test on the dominant hand of four healthy volunteers (3 males and 1 female, aged 24–27 years, with a mean age of 26.5 ± 1.7 years). The same algometer was used. Three sessions were conducted on three different days to assess PDTs during baseline, sham, and FUS conditions at the same three anatomical points. On days 2 and 3, the median nerve was identified, and each participant wore the fully wearable device. Counterbalancing was applied as above.

### Cold Pressor Task (CPT) Test

The second test was conducted over three sessions on different days, with eight healthy subjects (5 males and 3 females, aged 21-39, mean 26.1 ± 4.4 years) **(Fig. 3j)**. Styrofoam ice chests were filled with water and ice; water temperature was maintained at 1–2 °C using a digital thermometer with ice added as needed. On day 1, participants submerged their dominant hand in ice water and indicated the moment they first perceived pain; elapsed time was recorded with a stopwatch as PDT. After each PDT, participants waited 5 min before re-submerging their hands; 12 repetitions were performed to establish individual baselines. On days 2 and 3, the median nerve was localized at the antecubital fossa (human atlas and Philips Lumify, Philips Medical, Eindhoven, the Netherlands) **(Supplementary Fig. S8)**, a transducer was placed on the forearm, and either sham or LIFU stimulation was applied for 5 min (randomized order). Post-stimulation PDTs were recorded 12 times at 5-min intervals as on day 1.

In the seventh test, *PerNeUS* was used to measure PDTs using the CPT test on four healthy volunteers (2 males and 2 females, aged 24–39 years, mean 29.25 ± 6.6 years). The same three-session protocol was followed for baseline, sham, and LIFU. On days 2 and 3, the median nerve was identified prior to applying the fully wearable device. Counterbalancing was applied as above.

### Extended LIFU Effects on Pain Thresholds

We sought to investigate the duration of the effect of LIFU neurostimulation. Third and fourth tests were conducted, one using PA and the other using CPT, to evaluate the duration of the post-stimulation effect.

The PA test was conducted over two sessions with four healthy volunteers (3 males and 1 female, aged 22–39 years, mean 29 ± 7.7 years) on separate days. On day 1, baseline PDTs were measured at the wrist, palm, and middle finger over 9 h, recorded every 1 h (**Fig. 4a**). On day 2, the median nerve was stimulated with LIFU for 5 min, followed by algometry every 1 h for 9 h.

The CPT test was conducted over two days with four healthy volunteers (3 males and 1 female, aged 24–39 years, mean 33.5 ± 6.8 years). On day 1, PDT was measured with the hand immersed in ice water. On day 2, LIFU stimulation (5 min) was applied to the median nerve, and CPT measurements were recorded every 1 h for 9 h.

### Acoustic hydrogel durability test

The fifth test focused on analyzing the durability of the acoustic hydrogel during LIFU stimulation tests over 21 days. Prior work reported stability up to 28 days ^24^. We conducted tests with three volunteers (2 male and 1 female, aged 21-23, mean 22 ± 1 years) in 7 sessions, every 3 days **(Fig. 4d)**. Each session included three phases: (i) baseline PDT by PA (40 min, 10-min intervals), (ii) 5- min LIFU, and (iii) post-stimulation PDT (40 min, 10-min intervals). The median nerve was located by ultrasound before stimulation; the wearable transducer was connected to the commercial ultrasound system for 5-min sonication.

### Statistical Analysis

All statistical analyses were performed using GraphPad Prism (v10.4). For the PA and CPT tests with the commercial system (n = 8 each) and the fully wearable device (n = 4 each), the Friedman test was used to analyze three data groups (baseline, sham, and LIFU). Multiple Post Hoc comparisons were conducted using Dunn’s test (baseline vs. sham, sham vs. LIFU, baseline vs. LIFU). A two-sided p < 0.05 was considered statistically significant.

For the duration experiments (PA and CPT, n = 4 each), the Wilcoxon matched-pairs signed-rank test compared baseline and LIFU conditions. For the long-term stability test (n = 3 participants, 7 time points), the Friedman test analyzed repeated measures, with Dunn’s post hoc comparisons between baseline and post-LIFU at each session.

The comparison between the commercial system and the fully wearable device was performed using the Kruskal–Wallis test with multiple comparisons (**Supplementary Fig. S13**). Groups were analyzed independently (unmatched) due to different sample sizes (commercial, n = 8; wearable, n = 4). Comparisons were made across identical conditions (baseline, sham, LIFU) and cross-condition contrasts (sham_commercial vs. LIFU_wearable; sham_wearable vs. LIFU_commercial). Significance was set at two-sided p < 0.05.

### Study approval

All experiments were performed in compliance with the approved University of Texas at Austin Institutional Review Board (STUDY00005942). The study was registered on ClinicalTrials.gov (NCT07160049).

### Data availability

Data is available upon request from the authors.

## Data Availability

All data produced in the present study are available upon reasonable request to the authors

## Acknowledgements

H. W. would like to acknowledge support from the Alzheimer’s Association New to the Field (AARG-NTF) research grant, Defense Advanced Research Projects Agency (DARPA) (REM-REST) grant, UT Austin Proof of Concept Award and Cockrell Innovation Grant and NIH Maximizing Investigators’ Research Award (MIRA) (R35) grant. J.J would like to acknowledge support from the Human Frontier Science Program (HFSP) Fellowship. We acknowledge BioRender.com for the figures drawing.

## Author Contributions

Conceptualization: W.D.M.B., Y.H., J.J., Q.C., T.W., X.Q., Y.J. and H.W.; Methodology: W.D.M.B., J.J., Q.C., T.W., D.W. and X.Q.; Software: Y.H. and T.W.; Validation: W.D.M.B. and Y.H.; Formal Analysis: W.D.M.B. and J.J; Investigation: W.D.M.B., J.J., T.W., K.W.K.T, MX.Y, MM.Y., L.G., and J.C.H.; Data Curation: W.D.M.B; Writing - Original Draft: W.D.M.B.,Y.H. and J.J.; Writing - Review & Editing: W.D.M.B, Y.H., J.J., K.W.K.T, MX.Y., MM.Y., J.C.H., X.Q., Q.C., D.W., Y.J. and H.W.; Visualization: W.D.M.B.,Y.H. and J.J.; Project Administration: W.D.M.B and H.W.; Supervision: X.Q., Y.J. and H.W.; Funding: H.W.

