## Supporting Information for "Fully Wearable Armband for Long-Term Peripheral Ultrasound Neuromodulation in Pain Management"

### AUTHOR INFORMATION

William D. Moscoso-Barrera[1,†], Yiming Han[2,†], Jinmo Jeong[1,†], QiLiang Chen[3,†], Thomas Wynn[1], Mengxia Yu[1], Kai Wing Kevin Tang[1], Mengmeng Yao[1], Ju-Chun Hsieh[1], David Wu[3], Landon Gauthreaux[4], Xiang Qian[3*], Yaoyao Jia[2*], Huiliang Wang[1]*

### AFFILIATION

[1] *Department of Biomedical Engineering, Cockrell School of Engineering, The University of Texas at Austin, Austin, Texas 78712, United States.*

[2] *Chandra Family Department of Electrical and Computer Engineering, Cockrell School of Engineering, The University of Texas at Austin, Austin, Texas 78712, United States.*

[3] *Stanford Medicine, Stanford University, Stanford, California, 94305, United States.*

[4] *Engineering, Austin Community College, Austin, Texas 78752, United States.*

[†]These authors contributed equally to this work.

*Corresponding to:

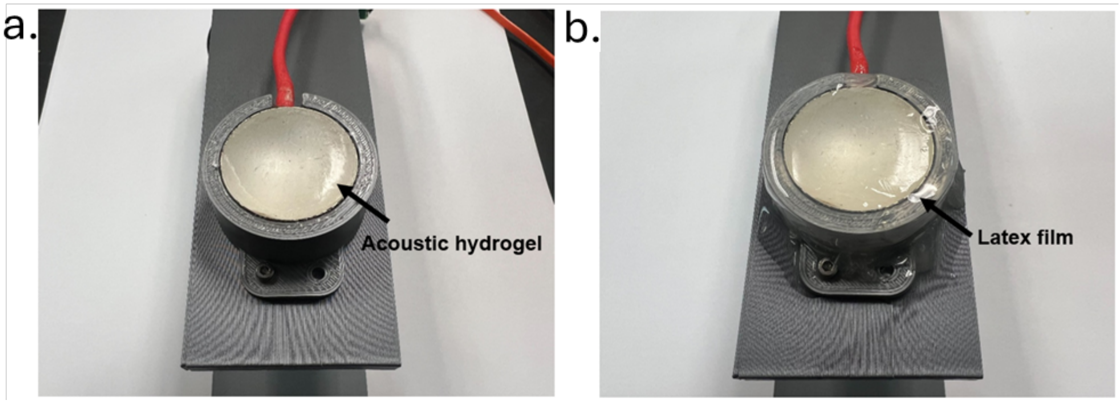

**Figure S1. CUT integrated with acoustic hydrogel for acoustic characterization in a water tank.**

**(a).** CUT integrated with acoustic hydrogel. **(b)** CUT with acoustic hydrogel wrapped in a thin latex film to prevent swelling during water-tank testing. The latex layer prevents hydrogel hydration and deformation during acoustic characterization.

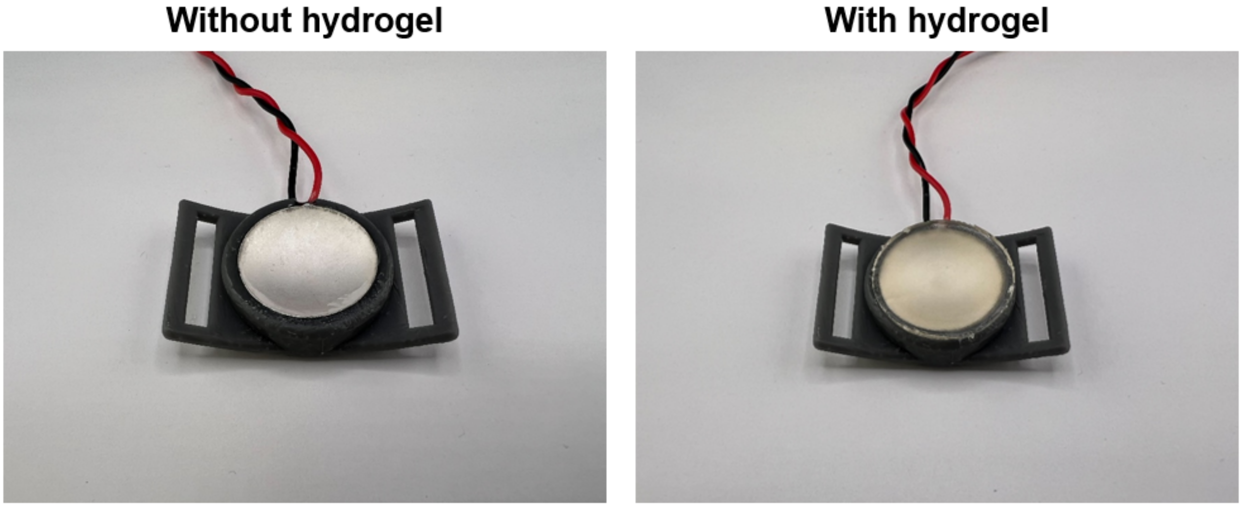

**Figure S2. CUT integrated with acoustic hydrogel mounted within the wearable armband housing.**

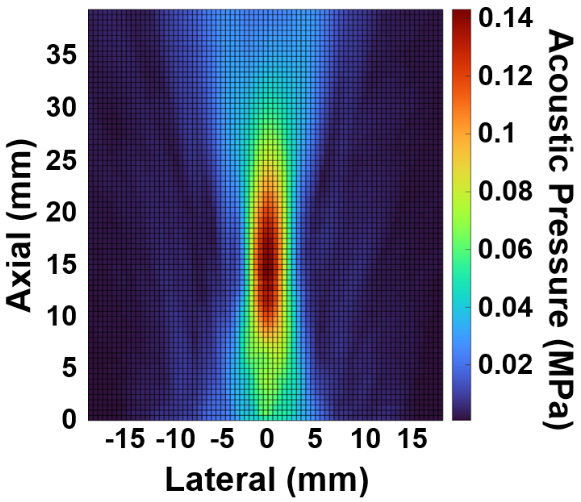

**Figure S3. Acoustic field distribution of the bare CUT without hydrogel integration.**

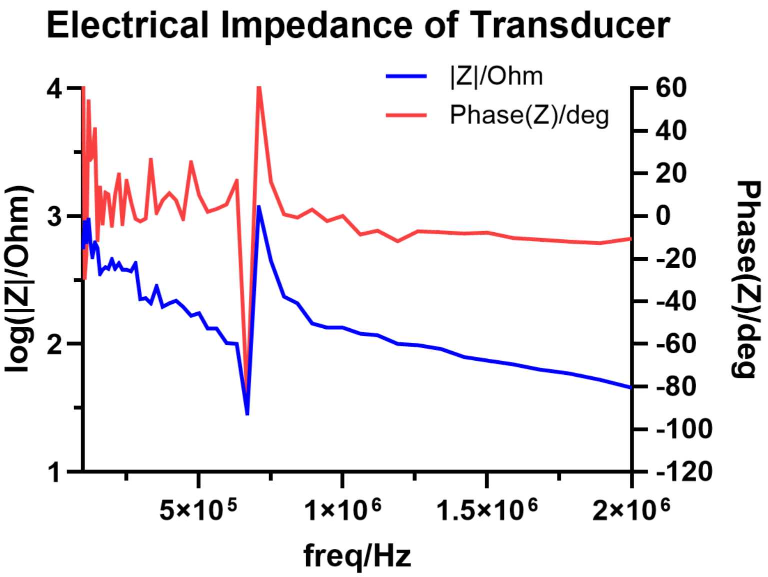

**Figure S4. Electrical impedance spectrum of the piezoelectric material integrated into the wearable transducer.**

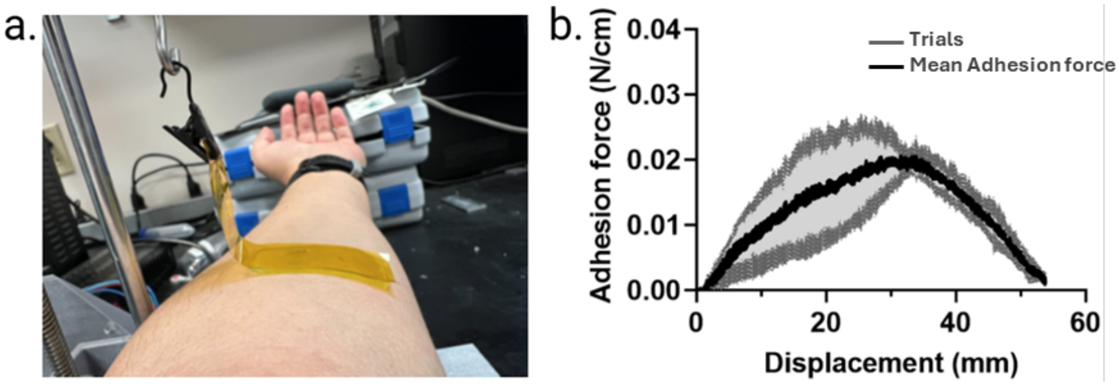

**Figure S5. Adhesion force test. (a)** Experimental setup for adhesion force measurement. **(b)** Adhesion force–displacement curves of the commercial ultrasonic gel (Aquasonic 100, Parker) on human skin (n = 3), with a magnified Y-axis view.The black curve represents the mean adhesion force across three trials, while the gray shaded region indicates the standard deviation, reflecting intertrial variability.

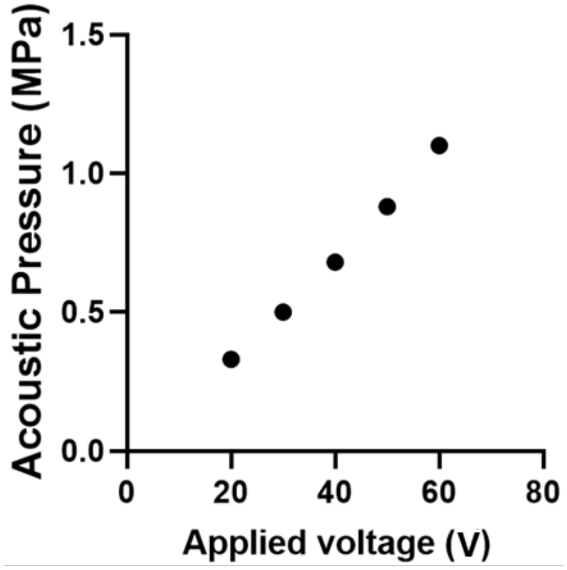

**Figure S6. Acoustic pressure generated by the CUT at different input voltages applied via the Vantage 64 LE system.**

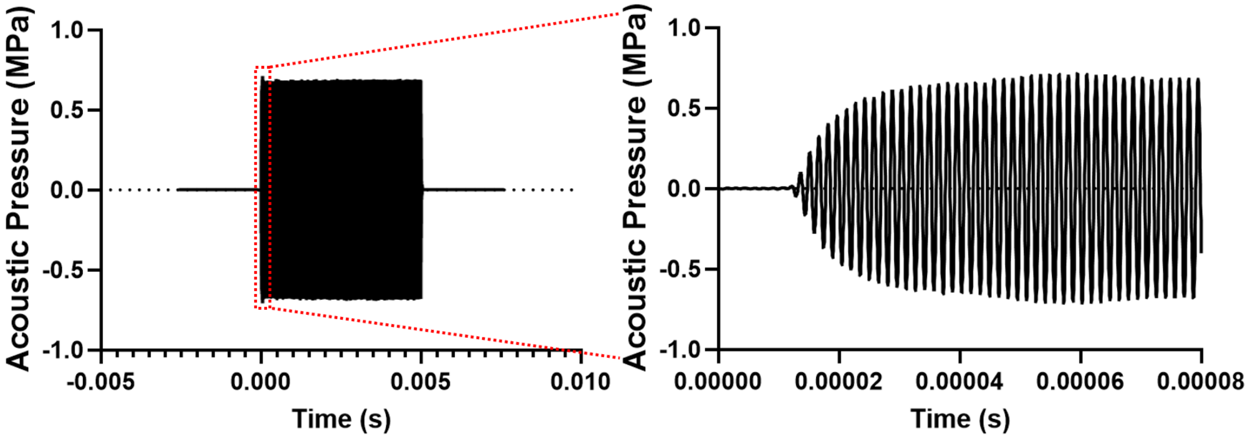

**Figure S7. Acoustic pressure waveform of a 5 ms ultrasound pulse generated by the CUT.**

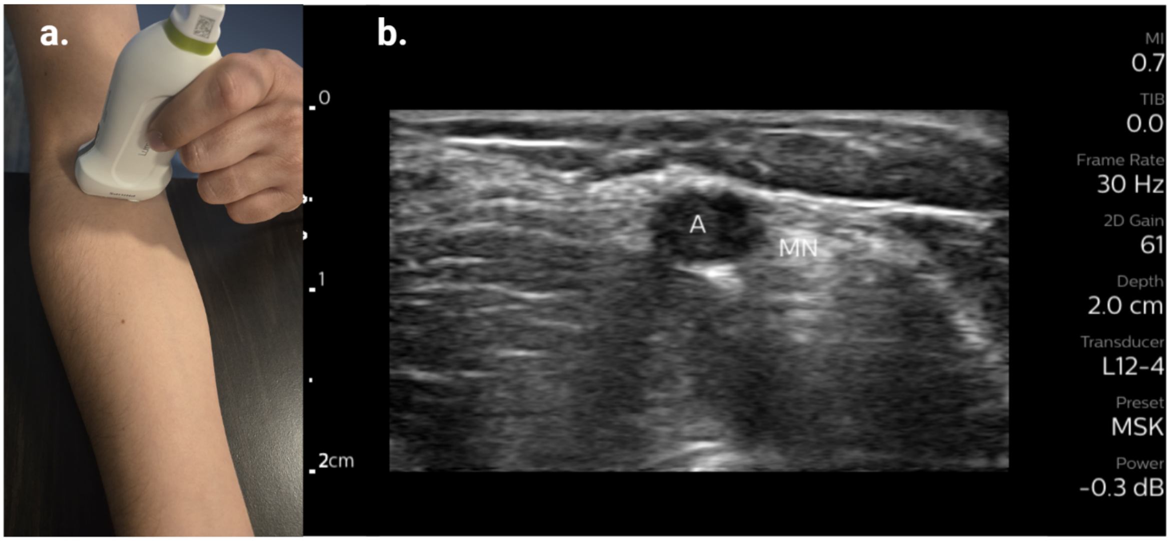

**Figure S8. Localization of the median nerve using ultrasound imaging.**

**(a)** Transducer positioning on the arm. **(b)** Cross-sectional ultrasound image showing the median nerve and the adjacent brachial artery (A).

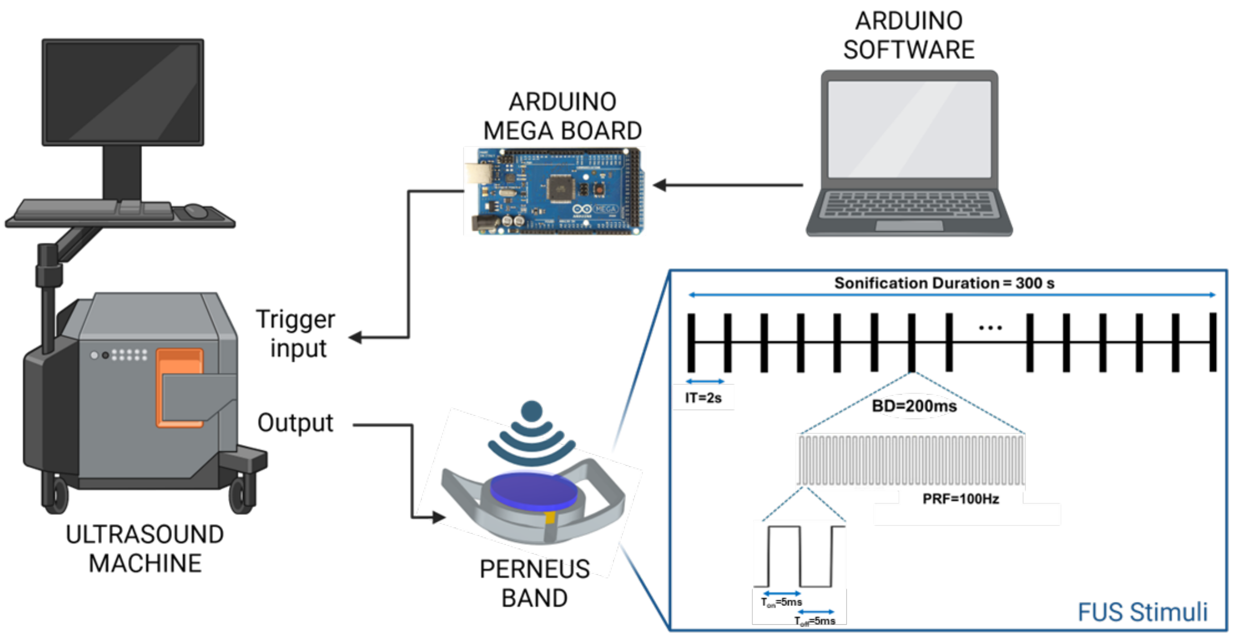

**Figure S9. Experimental setup for transmitting signals from the commercial ultrasound control system to the wearable transducer.**

Abbreviations: Sonification Duration (SD), Interpulse Time (IT), Burst Duration (BD), Pulse Repetition Frequency PRF, Time on ($T_{on}$), Time Off ($T_{off}$).

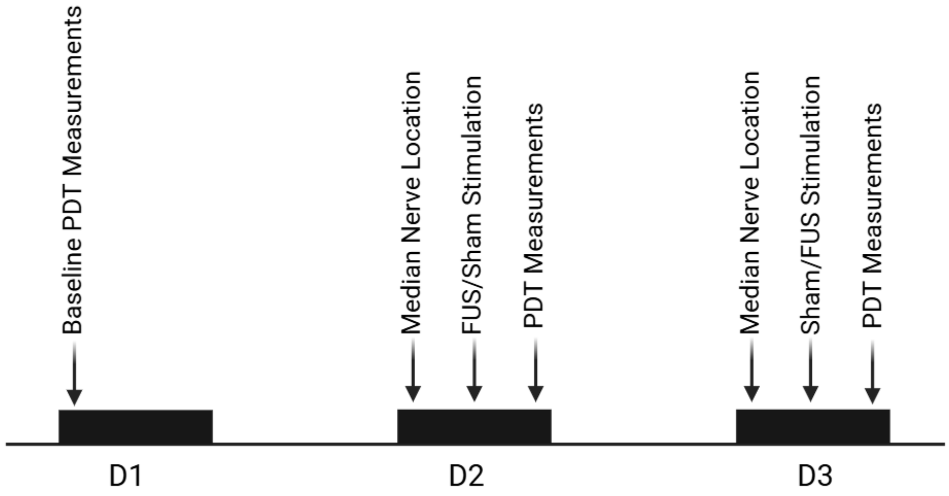

**Figure S10. Timeline of the Pressure Algometry (PA) Test.**

On Day 1 (D1), baseline pain detection thresholds (PDTs) were recorded. On Days 2 (D2) and 3 (D3), the median nerve was localized using ultrasound imaging, followed by either focused ultrasound stimulation (FUS) or sham stimulation using the wearable device, after which PDTs were remeasured.

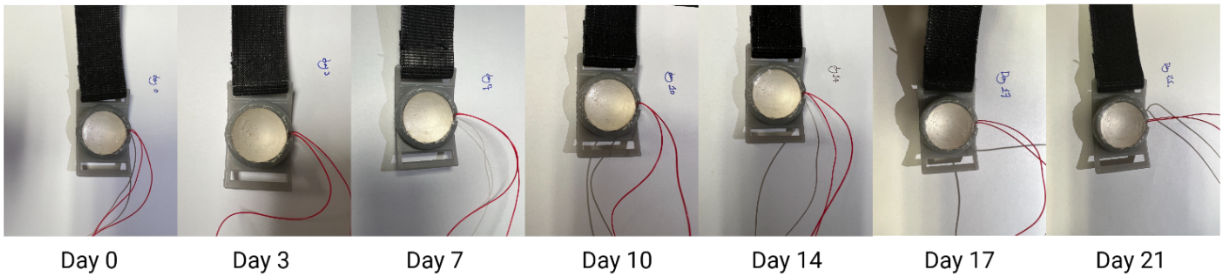

**Figure S11. Stability of the acoustic hydrogel over 21 days of continuous testing.** Images show the hydrogel maintaining structural integrity and adhesion throughout the testing period.

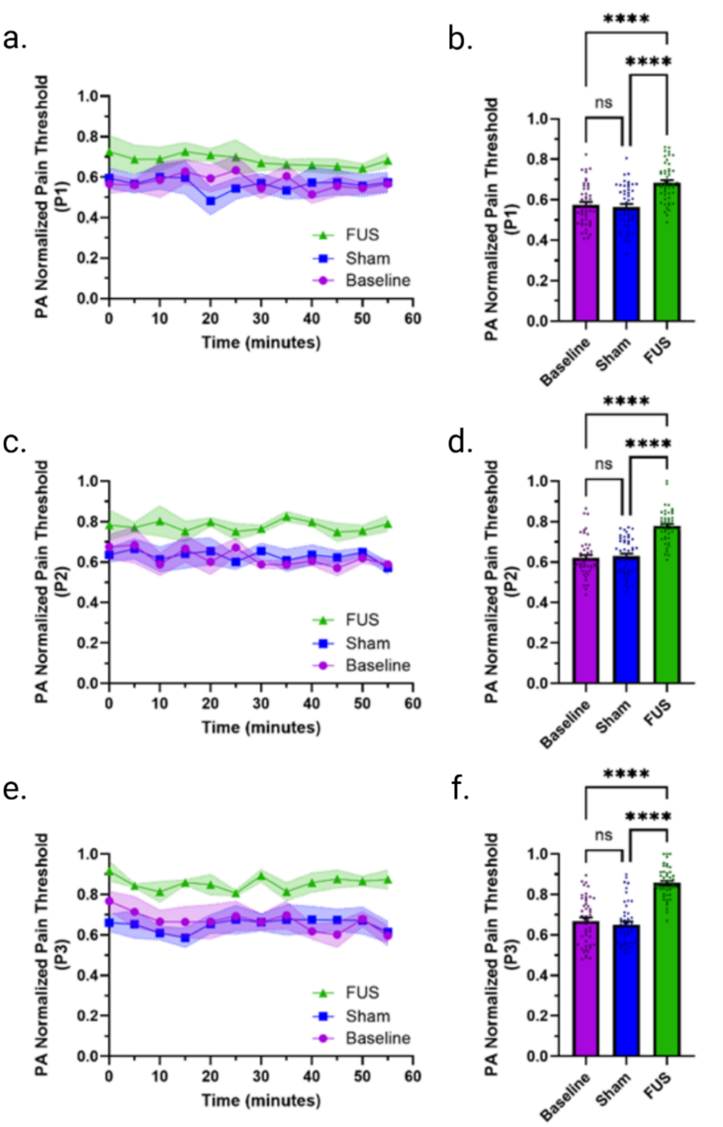

**Figure S12. Pain detection threshold tests using Pressure Algometry (PA) with the fully wearable system.**

**(a, c, e)** Time-series plots of aggregated normalized PDTs for sites P1, P2, and P3. **(b, d, f)** PA Aggregated normalized PDTs for sites P1, P2, and P3 under baseline, sham, and FUS conditions.

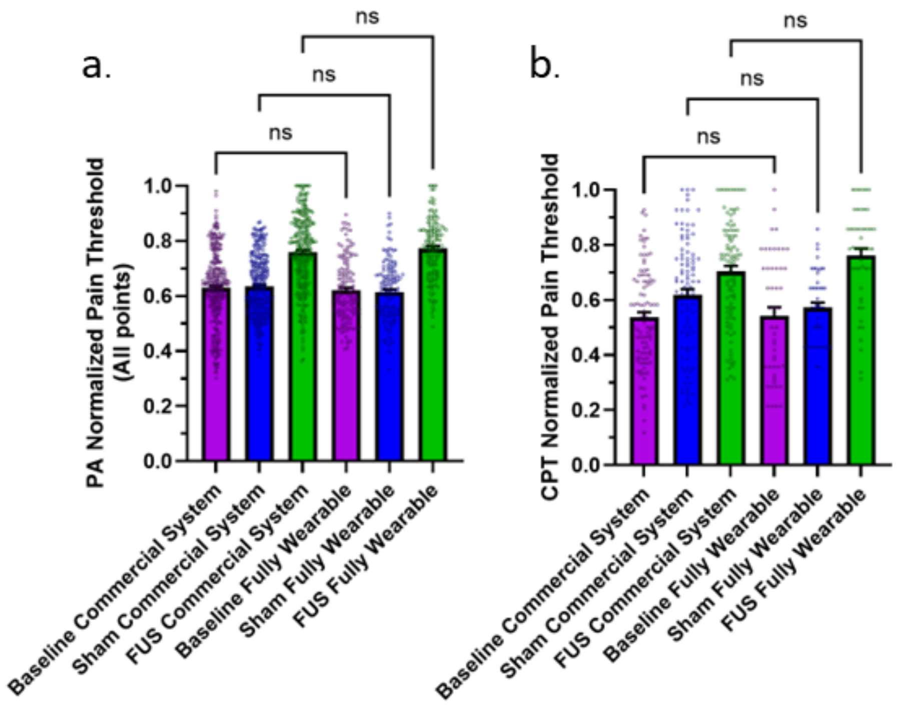

**Figure S13. Comparison between a commercial ultrasound stimulation system and the fully wearable *PerNeUS* device.**

**(a)** Comparison of PA test results obtained with the commercial ultrasound stimulation system and the fully wearable device. **(b)** Comparison of CPT test results obtained with the commercial ultrasound stimulation system and the fully wearable device. In experiments using the commercial system, the stimulator was connected to the wearable transducer. The fully wearable *PerNeUS* device integrates both the wearable transducer and the miniaturized electronic circuit.
